# Therapeutic use of convalescent plasma in COVID-19 patients with immunodeficiency

**DOI:** 10.1101/2020.11.08.20224790

**Authors:** Jonathon W. Senefeld, Stephen A. Klassen, Shane K. Ford, Chad C. Wiggins, Bruce C. Bostrom, Michael A. Thompson, Sarah E. Baker, Wayne T. Nicholson, Patrick W. Johnson, Rickey E. Carter, Jeffrey P. Henderson, William R. Hartman, Liise-anne Pirofski, R. Scott Wright, DeLisa Fairweather, Katelyn A. Bruno, Nigel S. Paneth, Arturo Casadevall, Michael J. Joyner

## Abstract

In the absence of effective countermeasures, human convalescent plasma has been widely used to treat severe acute respiratory syndrome coronavirus 2 including among patients with innate or acquired immunodeficiency. However, the association between COVID-19-associated mortality in patients with immunodeficiency and therapeutic use of convalescent plasma is unknown. We review clinical features and treatment protocols of COVID-19 patients with immunodeficiency after treatment with human convalescent plasma. We also discuss the time course and clinical features of recovery. These insights provide evidence for the need to develop a clear treatment protocol for COVID-19 patients with immunodeficiency and support the efficacy of convalescent plasma in patients with primary or secondary immunodeficiency.

## 1 Introduction

Convalescent plasma represents a passive antibody therapy that has been used to prevent or treat infectious diseases for more than a century.^1,2^ In the context of the novel coronavirus disease 2019 (COVID-19) pandemic, convalescent plasma has received full or conditional regulatory authorization in the United States for therapeutic use in adults and children hospitalized with suspected or laboratory confirmed SARS-CoV-2 positive COVID-19.^3–5^ Although early signals for efficacy and optimal use of convalescent plasma have emerged,^6–10^ additional large randomized controlled trials are needed to garner clinical consensus regarding the therapeutic effectiveness of convalescent plasma.

Inasmuch as convalescent plasma confers antiviral properties for the recipient, evidence of therapeutic effectiveness may be confounded by endogenous anti-SARS-CoV-2 antibody responses, particularly when convalescent plasma transfusion occurs late in the disease course.^11^ Hence, evaluation of the clinical responses to convalescent plasma transfusion in immunosuppressed patients who cannot generate innate immune responses may provide an optimal opportunity to assess the effect of convalescent plasma *per se*.^12^ Thus, herein we summarize the literature detailing the clinical experiences of patients with primary and secondary immunosuppression who were transfused with COVID-19 convalescent plasma. We hypothesize that patients with immunodeficiency syndromes, or patients with diseases associated with blunted endogenous antibody responses to COVID-19, may provide evidence of improved clinical status when given plasma containing anti-SARS-CoV-2 antibodies, with improvements over and above expectation in light of their underlying clinical conditions and the severity of COVID-19.

This review highlights 40 reports including 104 COVID-19 patients with primary immunosuppression due to Agammaglobulinemia (X-linked or autosomal) or Common Variable Immunodeficiency, and secondary immunodeficiencies related to hematological malignancies and solid organ transplants who were transfused with convalescent plasma. We also highlight additional cases of patients with COVID-19, including but not limited to patients with other forms of immunosuppression, transfused with convalescent plasma. We provide a summary overview of patient characteristics, COVID-19 therapies used, including convalescent plasma, and clinical symptomology (**table**).

**Search strategy and selection criteria**

References for this Review were identified through searches of PubMed for articles published from Jan 1, 2020 to Nov 1, 2020, using the Medical Subject Headings terms “COVID-19”, “convalescent plasma”, “convalescent serum”, “immunosuppression”, “cancer”, “transplant”, “agammaglobulinemia”, “malignancy”, and any relevant entry terms and supplementary concepts (**figure**). Relevant articles and data were also identified through searches in Google Scholar, medR_Χ_iv, and other websites. Articles resulting from these searches and relevant references cited in those articles were reviewed. Articles published in English were identified and included. All procedures accessed public information and did not require ethical review as determined by the Mayo Clinic Institutional Review Board in accordance with the Code of Federal Regulations, 45 CFR 46.102, and the Declaration of Helsinki.

**Table:** Summary of each patient’s hospital course and disposition after transfusion of COVID-19 convalescent plasma.

| Condition | No. Patients | WHO Disease Severity Scale* | Illness onset to Treatment (days)* | Mortality (n, %) | Rapid Improvement in Supplemental Oxygen (≤5 days) (n, %)* | Discharge (days)* |
| --- | --- | --- | --- | --- | --- | --- |
| **Primary Immunosuppression** |  |  |  |  |  |  |
| Agammaglobulinemia | 7 | 3.4 (2 - 5) | 26.3 (16 - 44) | 0, 0% | 2, 50% | 4.3 (1-7) |
| Common Variable Immune Deficiency | 4 | 2.8 (2 - 5) | .. | 1, 25% | .. | .. |
| **Secondary Immunosuppression** |  |  |  |  |  |  |
| Hematological Malignancies | 54 | 3.2 (2 - 5) | 23.8 (2 - 90) | 7, 13.0% | 22, 61.1% | 17.1 (2 - 50) |
| Solid Organ Transplants | 29 | 3.9 (2 - 5) | 10.6 (2 - 31) | 4, 13.8% | 14, 56.0% | 14.0 (2 - 58) |
| **Other<sup>a</sup>** | 10 | 4.0 (3 - 5) | 14.3 (6 - 39) | 1, 10.0% | 4, 40.0% | 18.1 (1.5 - 60) |

Data are presented as mean (range) or count (n), mean (%). \*Data were not reported for all patients, please see Supplement 1 for additional information. <sup>a</sup>Other includes pediatric patients, patients with congenital heart disease, gravida patients, and patients with Sjögren's syndrome

**Figure:**
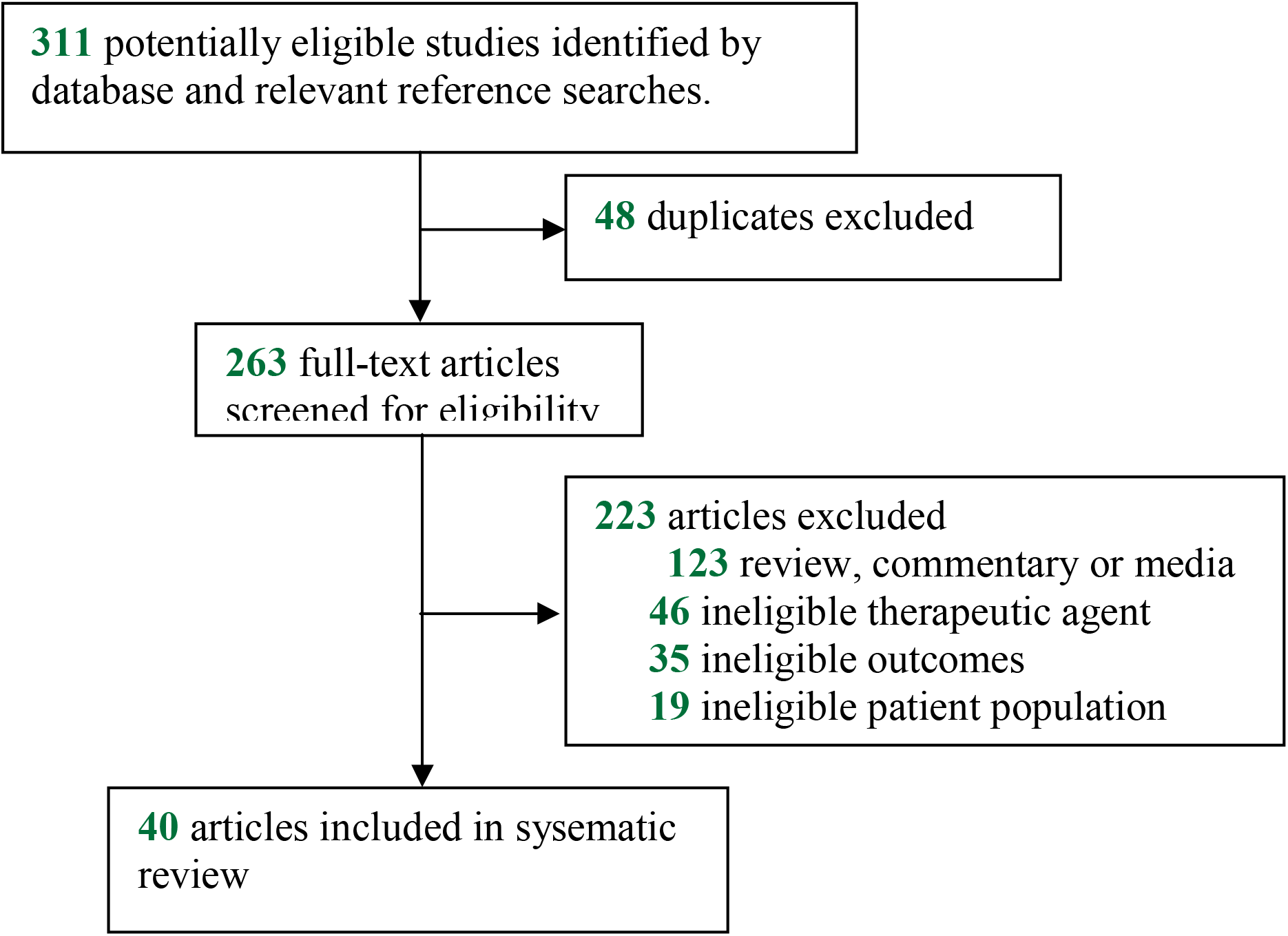
Flowchart of the study selection.

## 2 Primary Immunodeficiency

### 2.1 Patients with Agammaglobulinemia

This narrative includes one patient with Autosomal Agammaglobulinemia and six patients with X-linked Agammaglobulinemia (XLA), six of whom were reported in peer-reviewed articles and one patient who was identified in a media report.^13–16^ Patients with Agammaglobulinemia do not produce endogenous antibodies and require regular intravenous infusions (monthly) or subcutaneous injections (bi-weekly) of immunoglobulins to avoid serial infections from various common pathogens.^17^ However, immunoglobulin replacement therapy cannot protect patients against pathogens for which antibodies are uncommon or absent in the immunoglobulin donor pool, such as the SARS-CoV-2 virus.^17^ The clinical narratives describing patients with Agammaglobulinemias all demonstrate a prolonged course of disease and absent antibody response to COVID-19. However, all patients with Agammaglobulinemia demonstrated clinical improvement and symptom resolution following convalescent plasma transfusion, with three patients improving rapidly and demonstrating SARS-CoV-2 serum antibodies after transfusion. In addition to immunoglobulin therapy, some patients also received the experimental COVID-19 therapies remdesivir and hydroxychloroquine.

### 2.2 Patients with Common Variable Immunodeficiency

We identified four patients with Common Variable Immunodeficiency transfused with convalescent plasma for COVID-19 therapy.^16^ Common Variable Immunodeficiency represents a heterogeneous collection of immunodeficiencies commonly characterized by intrinsic B-cell defects and suppressed antibody production.^18^ Patients with Common Variable Immunodeficiency often present with inflammatory and autoimmune disorders, which are suspected to elevate these patients risk for progression to severe COVID-19.^16,18^ The four patients described here were all antibody deficient and diagnosed with severe or life-threatening COVID-19. Notably, three of these patients survived following convalescent plasma transfusion, including two patients whose clinical symptomatology required mechanical ventilation or extracorporeal membrane oxygenation.

## 3 Secondary Immunodeficiency

### 3.1 Patients with Hematological Malignancies

We identified 54 patients with hematological malignancies transfused with convalescent plasma in 18 peer-reviewed reports.^19,20,28,29,29–36,21–24,24–27^ The largest cohort of patients with hematological malignancies was a group of 17 patients with B-cell depletion whose immunodeficiency was secondary to therapies for various types of lymphoma or leukemia.^19^ These patients experienced protracted severe COVID-19 symptoms and absent endogenous SARS-CoV-2 antibody responses due to cancer-directed therapies that suppress B-cell proliferation, such as rituximab. Within 48 hours of convalescent plasma transfusion the majority of patients demonstrated improved clinical status and viral clearance. Similarly, in a separate cohort of 14 patients with hematological malignancies, most patients exhibited improvements in clinical symptomatology, including reduced oxygen requirements,^20^ after treatment with convalescent plasma. Other case-reports are of lymphoma (*n* = 11), leukemia (*n* = 5), multiple myeloma (*n* = 3), and myelodysplastic syndrome (*n* = 1) and include one four year old pediatric patient with leukemia who received two convalescent plasma doses over two days.^28^ A majority of patients with hematological malignancies recovered following convalescent plasma transfusion, with many demonstrating rapid clinical improvements shortly after transfusion. Notably, a patient with protracted COVID-19, evidenced by three separate COVID-19-related hospitalisations over a 100+ day period, and with lymphoma-associated B-cell immunodeficiency demonstrated rapid reductions in fever, oxygen requirements, and lung infiltrates (via chest computed tomography) forthwith after two separate convalescent plasma transfusions separated by ∼90 days.^22^

### 3.2 Solid Organ Transplant Patients

Among nine peer-reviewed articles and two media reports we identified a total of 29 COVID-19 patients transfused with convalescent plasma whom were receiving immunosuppressive therapies for previous solid organ transplants.^37–47^ In a cohort of 13 transplant recipients transfused with convalescent plasma concomitant to hydroxychloroquine, steroids and anticoagulants therapies for COVID-19, eight patients demonstrated improved oxygen requirements and were promptly discharged ^37^. In the other nine reports, a majority of the 17 patients with COVID-19 demonstrated improved clinical symptomatology following convalescent plasma transfusion. Improvement was even seen in a liver transplant patient discharged home after a convalescent plasma transfusion that occurred during a seventeen day medically-induced coma due to COVID-19 complications.^44^

## 4 Conclusion

These data provide evidence and encouraging anecdotal statements (**panel**) supporting the efficacy of convalescent plasma in patients with primary or secondary immunodeficiency, and are consistent with the historical evidence demonstrating that passive antibody therapies for infectious diseases are especially effective when given early in the course of disease prior to an endogenous antibody response.^2,48,49^ In contrast to ongoing studies of convalescent plasma efficacy in clinical trials where the majority of patients are not immunosuppressed and thus mount their own protective antibody responses,^6,11,50^ convalescent plasma use in these immunosuppressed patients represents a situation where exogenous antibody is provided in the setting of an immune deficit.^17^ This is particularly true in the case for patients with Agammaglobulinemia, who represent in effect a natural ‘immunological knockout’ providing the opportunity to evaluate the value of adding specific antibody to a host unable to make antibody to SARS-CoV-2.

The results for patients with Agammaglobulinemia are striking for the beneficial effects of convalescent plasma, and even though the reports lack a control group, the rapid time course of clinical improvement following administration of specific antibody in a situation where no endogenous antibody was present is impressive. Convalescent plasma was also found to be safe and efficacious in COVID-19 patients with common variable immunodeficiency, myasthenia gravis, and Sjögren’s syndrome, in addition to COVID-19 patients with heart rhythm abnormalities and gravida.^51–57^ Hence, from the viewpoint of establishing the efficacy of convalescent plasma administration in COVID-19 therapy, the experience with this patient set provides the important criterion that addition of specific antibody to a host with no antibodies resulted in a favorable therapeutic effect.

## Data Availability

These data are publicly available in the primary literature in which these data were abstracted.

## Contributors

MJJ is the guarantor. SAK, JWS, SKF, AC and MJJ conceptualised the paper, extracted the data and established the writing consortium. All authors contributed to the reviewing and editing of the report and approved the final version.

## Acknowledgements

We thank the members of the National COVID-19 Convalescent Plasma Project (http://ccpp19.org) for their support and collation of key scientific papers.

**Panel :Anecdotal statements supporting the efficacy of convalescent plasma**

- In the present case, the rapid clinical improvement followed by viral clearance after administration of hyperimmune plasma argue that passively transferred antibodies played a key role in COVID-19 recovery.^23^
- One day later [after convalescent plasma transfusion], the patient was afebrile for the first time in 3 weeks and had improved energy.^13^
- On day 122 (of illness), due to worsening symptoms, the patient was given convalescent plasma. He defervesced within 24 hours and was discharged nine days later.^31^
- …she was transferred to the intensive care unit for intubation. In the meantime…the patient received convalescent therapy instead and did not undergo intubation following the immediate improvement after plasma therapy infusion.^52^
- Based on the lack of clinical improvement…we transfused 1 unit of convalescent plasma…Importantly, the patient did not receive any other treatment potentially having an effect on the course of COVID-19…After transfusion of the convalescent plasma, the patient showed a dramatic clinical improvement, became asymptomatic, and was discharged home only 2 days later.^36^
- The patient was discharged after 2 weeks [convalescent plasma transfusion] with a dramatic response to therapy. Both newborns had no COVID-19 symptoms and negative PCR results.^54^
- 36 hours after [convalescent plasma] transfusion, the patient was discharged from the hospital reporting that he felt improved.^53^
- COVID-19 antibody testing showed complete lack of COVID-19 antibodies…She received 2 units of convalescent plasma…with rapid improvement in oxygen requirements. She was weaned off high-flow nasal cannula within 48 h and within a few days was discharged home in stable condition.^32^
- Intravenous convalescent plasma…was administered…Her health condition quickly improved, allowing [withdrawal of oxygen supplementation]…^23^
- Within a day of receiving her first transfusion of convalescent plasma, she reported improvement in shortness of breath and cough, had fever resolution, and decreasing oxygen requirements.^24^
- She received COVID convalescent plasma…She showed remarkable improvement [the next day]…with reduction of respiratory rate…and oxygen requirements.^21^
- …the patient received a transfusion of convalescent plasma…one day later her [arterial oxygen saturation] increased to 98%…clinical symptoms and pathological criteria improved rapidly within 3 days.^29^
- Within hours after receiving the convalescent plasma… [The patient’s] fever started going down. Days later, his breathing and kidney function improved.^45^
- [The patient]…received a transfusion of convalescent plasma…He is recovering at home after spending two and a half weeks in a coma fighting for his life…^44^
- …received 217 mL of convalescent plasma…24 hours later, his heart rate had improved to 60-70 bpm with less frequent premature atrial contractions and premature ventricular contractions and he was breathing comfortably on room air…36 hours after transfusion the patient was discharged from the hospital…^53^
- Stagnancy in the patient’s evolution, as represented by the lack of response to any of the treatments dispensed we administered on day 23 COVID-19 convalescent plasma. after 24 hours of infusion, fever ceased without subsequent reappearance and with progressive improvement of asthenia.^14^

## Supplementary Appendix

This appendix has been provided by the authors to give readers additional information about their work.

**Supplement to:** Therapeutic use of convalescent plasma in COVID-19 patients with immunodeficiency

**Most Recent Update:** November 8, 2020

**Supplementary Table 1:**
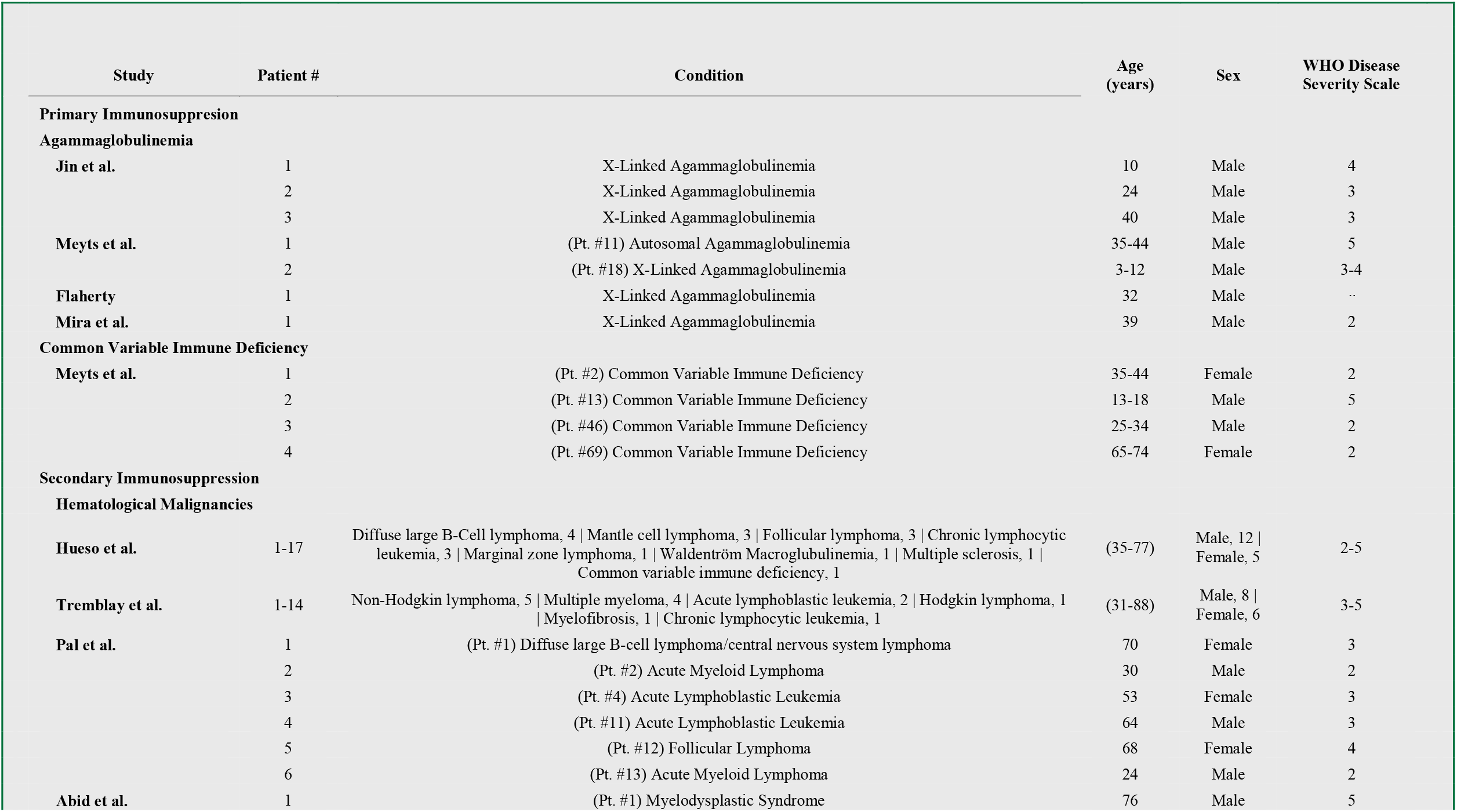
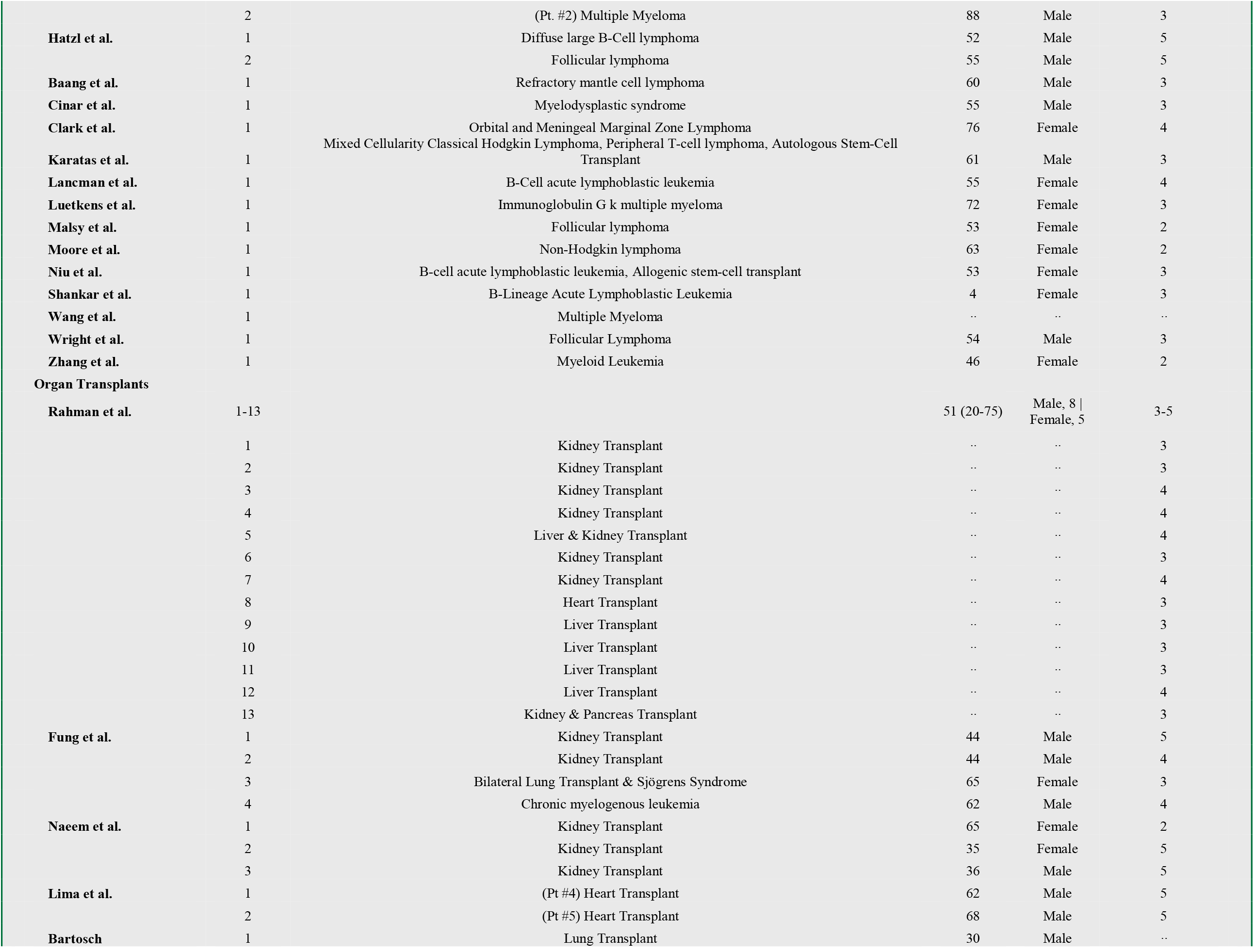
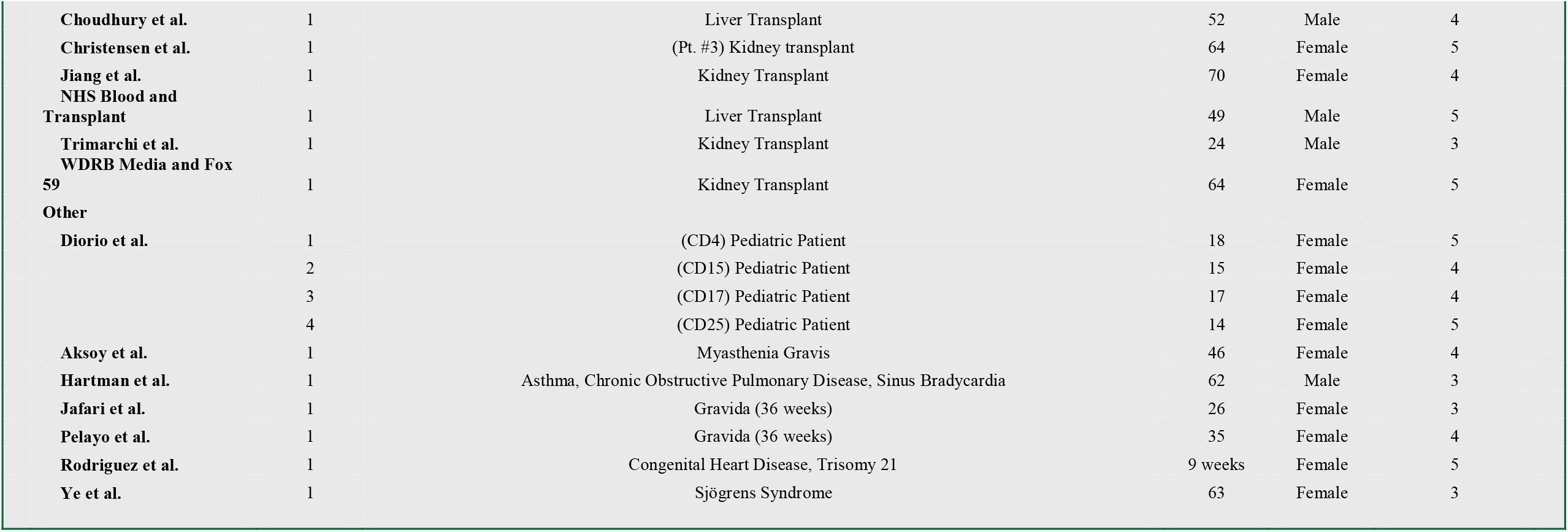
Summary of patient’s characteristics prior to transfusion of COVID-19 convalescent plasma.

**Supplementary Table 2:**
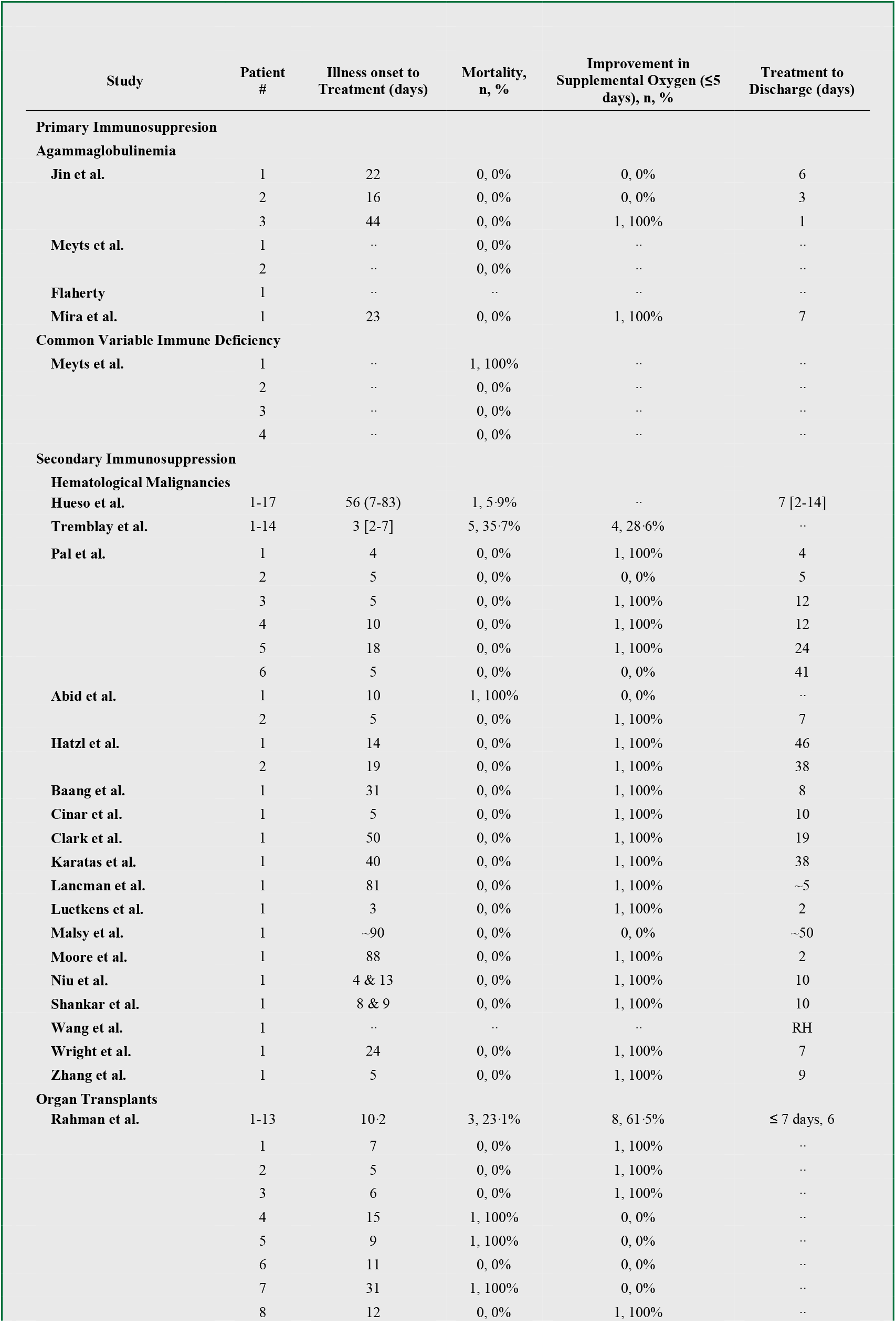
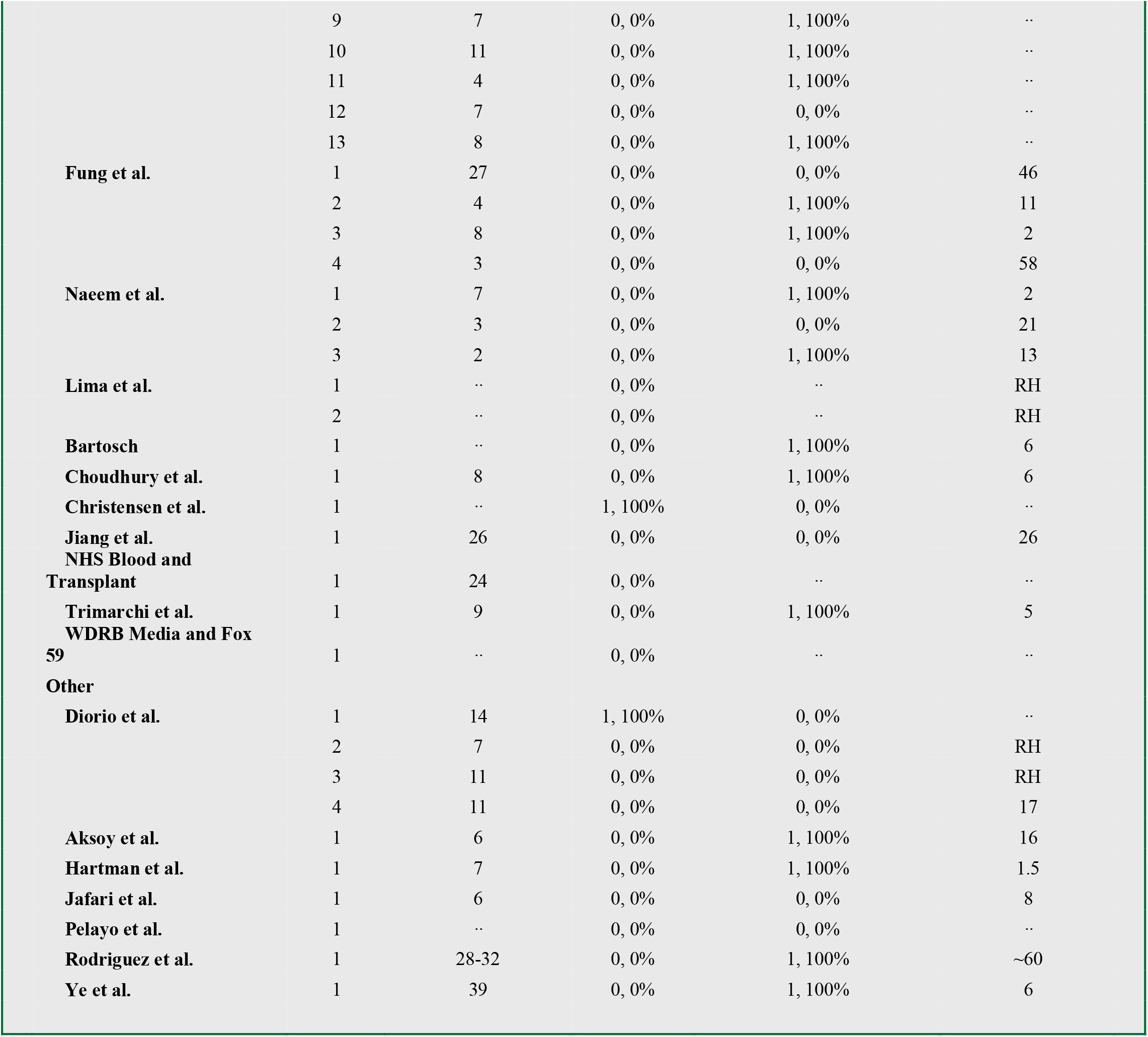
Summary of patient’s hospital course and disposition after transfusion of COVID-19 convalescent plasma.

**Supplementary Table 3:** Summary of patient’s concomitant COVID-19 therapeutics prior to transfusion of COVID-19 convalescent plasma.

| Study | Patient # | Concomitant therapies |  |  |  |  |
| --- | --- | --- | --- | --- | --- | --- |
|  |  | IVIG | Steroids | remdesivir | (hydroxy)chloroquine | antibiotics |
| Primary Immunosuppression |  |  |  |  |  |  |
| Agammaglobulinemia |  |  |  |  |  |  |
| Jin et al. | 1 | Y |  | Y |  |  |
|  | 2 | Y |  |  |  | Y |
|  | 3 |  |  |  |  | Y |
| Meyts et al. | 1 | Y |  |  | Y | Y |
|  | 2 |  |  | Y |  | Y |
| Flaherty | 1 |  |  | Y |  |  |
| Mira et al. | 1 | Y |  |  | Y | Y |
| Common Variable Immune Deficiency |  |  |  |  |  |  |
| Meyts et al. | 1 | Y | Y |  | Y | Y |
|  | 2 |  | Y | Y |  | Y |
|  | 3 | Y | Y |  | Y | Y |
|  | 4 |  |  |  | Y | Y |
| Secondary Immunosuppression |  |  |  |  |  |  |
| Hematological Malignancies |  |  |  |  |  |  |
| Hueso et al. | 1-17 |  | Y | Y | Y | Y |
| Tremblay et al. | 1-14 |  |  | Y | Y |  |
| Pal et al. | 1 |  |  | Y |  |  |
|  | 2 |  |  | Y |  |  |
|  | 3 |  |  |  |  |  |
|  | 4 |  |  |  |  |  |
|  | 5 |  |  | Y |  |  |
|  | 6 |  |  |  |  |  |
| Abid et al. | 1 |  |  | Y |  |  |
|  | 2 |  |  |  |  |  |
| Hatzl et al. | 1 |  | Y |  | Y | Y |
|  | 2 |  | Y |  | Y | Y |
| Baang et al. | 1 |  |  | Y |  |  |
| Cinar et al. | 1 |  |  |  |  |  |
| Clark et al. | 1 |  | Y |  |  |  |
| Karatas et al. | 1 |  |  |  | Y | Y |
| Lancman et al. | 1 |  | Y |  | Y | Y |
| Luetkens et al. | 1 |  | Y |  |  |  |
| Malsy et al. | 1 |  |  | Y |  |  |
| Moore et al. | 1 |  |  |  |  |  |
| Niu et al. | 1 | Y |  |  |  | Y |
| Shankar et al. | 1 | Y | Y |  |  | Y |
| Wang et al. | 1 | .. | .. | .. | .. | .. |
| Wright et al. | 1 |  |  |  | Y | Y |
| Zhang et al. | 1 |  |  |  |  | Y |
| Organ Transplants |  |  |  |  |  |  |
| Rahman et al. | 1-13 |  | Y |  | Y |  |
|  | 1 | .. | .. | .. | .. | .. |
|  | 2 | .. | .. | .. | .. | .. |
|  | 3 | .. | .. | .. | .. | .. |
|  | 4 | .. | .. | .. | .. | .. |
|  | 5 | .. | .. | .. | .. | .. |
|  | 6 | .. | .. | .. | .. | .. |
|  | 7 | .. | .. | .. | .. | .. |
|  | 8 | .. | .. | .. | .. | .. |
|  | 9 | .. | .. | .. | .. | .. |
|  | 10 | .. | .. | .. | .. | .. |
|  | 11 | .. | .. | .. | .. | .. |
|  | 12 | .. | .. | .. | .. | .. |
|  | 13 | .. | .. | .. | .. | .. |
| **Fung et al.** | 1 |  | Y |  | Y | Y |
|  | 2 |  |  |  |  |  |
|  | 3 |  |  | Y |  |  |
|  | 4 |  |  |  |  |  |
| **Naeem et al.** | 1 |  | Y |  |  |  |
|  | 2 |  | Y | Y |  | Y |
|  | 3 |  | Y | Y |  | Y |
| **Lima et al.** | 1 |  | Y |  |  |  |
|  | 2 |  | Y |  |  |  |
| **Bartosch** | 1 | .. | .. | .. | .. | .. |
| **Choudhury et al.** | 1 |  | Y | Y | Y | Y |
| **Christensen et al.** | 1 |  |  |  | Y |  |
| **Jiang et al.** | 1 |  | Y |  |  | Y |
| **NHS Blood and Transplant** | 1 | .. | .. | .. | .. | .. |
| **Trimarchi et al.** | 1 |  | Y |  |  | Y |
| **WDRB Media and Fox 59** | 1 | .. | .. | .. | .. | .. |
| **Other** |  |  |  |  |  |  |
| **Diorio et al.** | 1 |  | Y |  | Y |  |
|  | 2 |  |  | Y |  |  |
|  | 3 |  | Y | Y |  |  |
|  | 4 |  | Y |  |  |  |
| **Aksoy et al.** | 1 |  | Y |  | Y | Y |
| **Hartman et al.** | 1 |  | Y |  |  |  |
| **Jafari et al.** | 1 |  |  |  | Y | Y |
| **Pelayo et al.** | 1 |  | Y | Y |  |  |
| **Rodriguez et al.** | 1 |  |  | Y | Y |  |
| **Ye et al.** | 1 |  |  |  |  | Y |

## References

1 Casadevall A, Pirofski L. The convalescent sera option for containing COVID-19. J Clin Invest 2020; 130: 1545–8.

2 Luke TC, Kilbane EM, Jackson JL, Hoffman SL. Meta-analysis: convalescent blood products for Spanish influenza pneumonia: a future H5N1 treatment? Ann Intern Med 2006; 145: 599–609.

3 Bloch EM, Shoham S, Casadevall A, et al. Deployment of convalescent plasma for the prevention and treatment of COVID-19. J Clin Invest 2020.

4 US Food and Drug Administration. Recommendations for Investigational COVID-19 Convalescent Plasma. 2020. https://www.fda.gov/vaccines-blood-biologics/investigational-new-drug-ind-or-device-exemption-ide-process-cber/recommendations-investigational-covid-19-convalescent-plasma.

5 US Food and Drug Administration. Clinical Memorandum for the Emergency Use Authorization of COVID-19 Convalescent Plasma. 2020 https://www.fda.gov/media/141480/download.

6 Li L, Zhang W, Hu Y, et al. Effect of Convalescent Plasma Therapy on Time to Clinical Improvement in Patients With Severe and Life-threatening COVID-19: A Randomized Clinical Trial. JAMA 2020.

7 Abolghasemi H, Eshghi P, Cheraghali AM, et al. Clinical Efficacy of Convalescent Plasma for Treatment of COVID-19 Infections: Results of a Multicenter Clinical Study. Transfus Apher Sci 2020; : 102875.

8 Salazar E, Christensen PA, Graviss EA, et al. Early transfusion of a large cohort of COVID-19 patients with high titer anti-SARS-CoV-2 spike protein IgG convalescent plasma confirms a signal of significantly decreased mortality. medRxiv 2020.

9 Joyner MJ, Senefeld JW, Klassen SA, et al. Effect of convalescent plasma on mortality among hospitalized patients with COVID-19: initial three-month experience. MedRxiv 2020.

10 Joyner MJ, Bruno K, Klassen SA, et al. Safety Update: COVID-19 Convalescent Plasma in 20,000 Hospitalized Patients. Mayo Clin Proc 2020; 95.

11 Agarwal A, Mukherjee A, Kumar G, Chatterjee P, Bhatnagar T, Malhotra P. Convalescent plasma in the management of moderate covid-19 in adults in India: open label phase II multicentre randomised controlled trial (PLACID Trial). bmj 2020; 371.

12 Patel SY, Carbone J, Jolles S. The expanding field of secondary antibody deficiency: causes, diagnosis, and management. Front Immunol 2019; 10: 33.

13 Jin H, Reed JC, Liu STH, et al. Three patients with X-linked agammaglobulinemia hospitalized for COVID-19 improved with convalescent plasma. J Allergy Clin Immunol Pract 2020.

14 Mira E, Yarce OA, Ortega C, et al. Rapid recovery of a SARS-CoV-2–infected X-linked agammaglobulinemia patient after infusion of COVID-19 convalescent plasma. J Allergy Clin Immunol Pract 2020; 8: 2793–5.

15 Flaherty C. Man leaves hospital after three month battle with Covid-19. Impartial Report. 2020. https://www.impartialreporter.com/news/18599490.man-leaves-hospital-three-month-battle-covid-19/ (Accessed Oct 25, 2020).

16 Meyts I, Bucciol G, Quinti I, et al. Coronavirus Disease 2019 in patients with inborn errors of immunity: an international study. J Allergy Clin Immunol 2020.

17 Mazhar M, Waseem M. Agammaglobulinemia. In: StatPearls [Internet]. StatPearls Publishing, 2020.

18 Abbott JK, Gelfand EW. Common variable immunodeficiency: diagnosis, management, and treatment. Immunol Allergy Clin 2015; 35: 637–58.

19 Hueso T, Pouderoux C, Péré H, et al. Convalescent plasma therapy for B-cell depleted patients with protracted COVID-19 disease. Blood 2020.

20 Tremblay D, Seah C, Schneider T, et al. Convalescent Plasma for the Treatment of Severe COVID□19 Infection in Cancer Patients. Cancer Med 2020.

21 Hatzl S, Eisner F, Schilcher G, et al. Response to “COVID-19 in persons with haematological cancers”. Leukemia 2020; 34: 2265–70.

22 Baang JH, Smith C, Mirabelli C, et al. Prolonged SARS-CoV-2 replication in an immunocompromised patient. medRxiv 2020.

23 Lancman G, Mascarenhas J, Bar-Natan M. Severe COVID-19 virus reactivation following treatment for B cell acute lymphoblastic leukemia. J Hematol Oncol 2020; 13: 1–3.

24 Moore JL, Ganapathiraju P V, Kurtz CP, Wainscoat B. A 63-Year-Old Woman with a History of Non-Hodgkin Lymphoma with Persistent SARS-CoV-2 Infection Who Was Seronegative and Treated with Convalescent Plasma. Am J Case Rep 2020; 21: e927812.

25 Çınar OE, Sayınalp B, Karakulak EA, et al. Covalescent (immune) plasma treatment in a myelodysplastic COVID-19 patient with disseminated Tuberculosis. Transfus Apher Sci 2020; : 102821.

26 Karataş A, İnkaya AÇ, Demiroğlu H, et al. Prolonged viral shedding in a lymphoma patient with COVID-19 infection receiving convalescent plasma. Transfus Apher Sci 2020; : 102871.

27 Luetkens T, Metcalf R, Planelles V, et al. Successful transfer of anti–SARS-CoV-2 immunity using convalescent plasma in an MM patient with hypogammaglobulinemia and COVID-19. Blood Adv 2020; 4: 4864–8.

28 Shankar R, Radhakrishnan N, Dua S, et al. Convalescent plasma to aid in recovery of COVID-19 pneumonia in a child with acute lymphoblastic leukemia. Transfus Apher Sci 2020; : 102956.

29 Wright Z, Bersabe A, Eden R, Cap A. Successful use of COVID-19 convalescent plasma in a patient recently treated for follicular lymphoma. Clin Lymphoma Myeloma Leuk 2020.

30 Clark E, Guilpain P, Filip IL, et al. Convalescent plasma for persisting Covid□19 following therapeutic lymphocyte depletion: a report of rapid recovery. Br J Haematol 2020; 190: e154–6.

31 Niu A, McDougal A, Ning B, et al. COVID-19 in allogeneic stem cell transplant: high false-negative probability and role of CRISPR and convalescent plasma. Bone Marrow Transplant 2020; : 1–3.

32 Pal P, Ibrahim M, Niu A, et al. Safety and efficacy of COVID-19 convalescent plasma in severe pulmonary disease: A report of 17 patients. Transfus Med 2020.

33 Abid MB, Chhabra S, Buchan B, et al. Bronchoalveolar lavage-based COVID-19 testing in patients with cancer. Hematol Oncol Stem Cell Ther 2020.

34 Malsy J, Veletzky L, Heide J, et al. Sustained response after remdesivir and convalescent plasma therapy in a B-cell depleted patient with protracted COVID-19. Clin Infect Dis 2020.

35 Wang B, Van Oekelen O, Mouhieddine T, et al. A tertiary center experience of multiple myeloma patients with COVID-19: lessons learned and the path forward. medRxiv 2020.

36 Zhang LL, Liu Y, Guo YG, et al. Convalescent Plasma Rescued a Severe COVID-19 Patient with Chronic Myeloid Leukemia Blast Crisis and Myelofibrosis. Turkish J Haematol Off J Turkish Soc Haematol 2020.

37 Rahman F, Liu STH, Taimur S, et al. Treatment with convalescent plasma in solid organ transplant recipients with COVID□19: Experience at large transplant center in New York City. Clin Transplant 2020; : e14089.

38 Fung M, Nambiar A, Pandey S, et al. Treatment of Immunocompromised COVID□19 patients with Convalescent Plasma. Transpl Infect Dis 2020.

39 Trimarchi H, Gianserra R, Lampo M, Monkowski M, Lodolo J. Eculizumab, SARS-CoV-2 and atypical hemolytic uremic syndrome. 2020.

40 Naeem S, Gohh R, Bayliss G, et al. Successful recovery from COVID□19 in three kidney transplant recipients who received convalescent plasma therapy. Transpl Infect Dis 2020; : e13451.

41 Lima B, Gibson GT, Vullaganti S, et al. COVID□19 in recent heart transplant recipients: Clinicopathologic features and early outcomes. Transpl Infect Dis 2020; : e13382.

42 Jiang J, Miao Y, Zhao Y, et al. Convalescent plasma therapy: Helpful treatment of COVID□19 in a kidney transplant recipient presenting with severe clinical manifestations and complex complications. Clin Transplant 2020; 34: e14025.

43 WDRB Media and Fox 59. Plasma donation helps save Indiana man diagnosed with COVID-19. 2020. https://www.wdrb.com/news/coronavirus/plasma-donation-helps-save-indiana-man-diagnosed-with-covid-19/article_540bc9b2-ac1d-11ea-958d-abf6e203c27a.html.

44 NHS Blood and Transplant. One of the first COVID-19 convalescent plasma recipients supports donor appeal. Give Blood. 2020. https://www.blood.co.uk/news-and-campaigns/news-and-statements/one-of-the-first-covid-19-convalescent-plasma-recipients-supports-donor-appeal/.

45 Bartosch J. Lung transplant patient with COVID-19 recovers following plasma clinical trial. Univ. Chicago Med. 2020. https://www.uchicagomedicine.org/forefront/coronavirus-disease-covid-19/lung-transplant-patient-with-covid-19-recovers-following-plasma-clinical-trial.

46 Choudhury A, Reddy GS, Venishetty S, et al. COVID-19 in Liver Transplant Recipients-A Series with Successful Recovery. J Clin Transl Hepatol 2020; 8: 1–7.

47 Christensen J, Kumar D, Moinuddin I, et al. Coronavirus Disease 2019 Viremia, Serologies, and Clinical Course in a Case Series of Transplant Recipients. In: Transplantation proceedings. Elsevier, 2020.

48 Cheng Y, Wong R, Soo YOY, et al. Use of convalescent plasma therapy in SARS patients in Hong Kong. Eur J Clin Microbiol Infect Dis 2005; 24: 44–6.

49 Hung IFN, To KKW, Lee C-K, et al. Convalescent plasma treatment reduced mortality in patients with severe pandemic influenza A (H1N1) 2009 virus infection. Clin Infect Dis 2011; 52: 447–56.

50 Gharbharan A, Jordans CCE, GeurtsvanKessel C, et al. Convalescent Plasma for COVID-19. A randomized clinical trial. medRxiv 2020.

51 Diorio C, Anderson EM, McNerney KO, et al. Convalescent plasma for pediatric patients with SARS□CoV□2□associated acute respiratory distress syndrome. Pediatr Blood Cancer 2020; 67: e28693.

52 Aksoy E, Oztutgan T. COVID-19 presentation in association with myasthenia gravis: a case report and review of the literature. Case Rep Infect Dis 2020; 2020.

53 Hartman W, Hess AS, Connor JP. Hospitalized COVID-19 patients treated with Convalescent Plasma in a mid-size city in the midwest. medRxiv 2020.

54 Jafari R, Jonaidi-Jafari N, Dehghanpoor F, Saburi A. Convalescent plasma therapy in a pregnant COVID-19 patient with a dramatic clinical and imaging response: A case report. World J Radiol 2020; 12: 137.

55 Pelayo J, Pugliese G, Salacup G, et al. Severe COVID-19 in Third Trimester Pregnancy: Multidisciplinary Approach. Case Reports Crit Care 2020; 2020.

56 Rodriguez Z, Shane AL, Verkerke H, et al. COVID-19 convalescent plasma clears SARS-CoV-2 refractory to remdesivir in an infant with congenital heart disease. Blood Adv 2020; 4: 4278.

57 Ye M, Fu D, Ren Y, et al. Treatment with convalescent plasma for COVID□19 patients in Wuhan, China. J Med Virol 2020.

